# Delayed Transcallosal Conduction to the Lesioned Sensorimotor Cortex in Multiple Sclerosis: A combined TMS 7T-MRI Study

**DOI:** 10.64898/2026.03.20.26348903

**Authors:** Mads A.J. Madsen, Lasse Christiansen, Vanessa Wiggermann, Henrik Lundell, Jeppe Romme Christensen, Morten Blinkenberg, Finn Sellebjerg, Hartwig R. Siebner

## Abstract

**Background:** In multiple sclerosis (MS), demyelination and degeneration of transcallosal pathways impair interhemispheric communication. While white matter damage is well documented, the impact of cortical lesions on transcallosal conduction remains unclear.

**Objective:** To determine whether cortical lesions in the sensorimotor hand area (SM1□HAND) contribute to impaired transcallosal motor interaction using ultra□high□field MRI and transcranial magnetic stimulation (TMS).

**Methods:** Twenty healthy controls (HCs) and 38 MS patients underwent 7T structural and diffusion□weighted MRI. Structural scans were used to identify cortical lesions in SM1□HAND, while diffusion tensor imaging (DTI) quantified microstructural properties in the transcallosal tract connecting left and right SM1□HAND. Single□pulse TMS was delivered to each SM1□HAND during tonic first dorsal interosseous contraction to measure the ipsilateral silent period (iSP). Corticospinal conduction was measured with contralateral motor□evoked potentials (MEPs), while the iSP was used to compute transcallosal conduction time (TCT).

**Results:** Among MS patients, 41 of 76 hemispheres contained an SM1□HAND lesion. TCT was significantly prolonged in MS relative to HCs (P<0.001). In patients, cortical lesions delayed transcallosal conduction from the non□lesion□bearing to the lesion□bearing hemisphere (P=0.026). This direction-specific delay was associated with an intracortical lesion type (P<0.001), but not with DTI□derived microstructural measures (P>0.05).

**Conclusions:** The presence of cortical lesions in the sensorimotor cortex affects transcallosal inhibition between homologous sensorimotor regions in MS, slowing the build-up of inhibitory influence on the corticospinal output in the lesioned cortex. This delayed inhibitory buildl⍰up appears to be associated with an intracortical lesion type.

**Highlights:**

- Ipsilateral silent period reveals delayed transcallosal motor interaction in multiple sclerosis
- Cortical lesions in sensorimotor cortex delay the onset of transcallosal motor inhibition
- Delayed transcallosal inhibition is only present toward the lesioned cortex
- Intracortical lesions, not callosal microstructure, is linked to this directionl⍰specific delay

## 1. Introduction

Multiple sclerosis (MS) is a demyelinating and neurodegenerative disease of the central nervous system characterized by focal lesions and diffuse pathology affecting both white and grey matter (Reich et al., 2018). White matter lesions are readily detected with conventional 1.5T or 3T magnetic resonance imaging (MRI). Cortical pathology, however, particularly focal lesions, are underestimated in routine clinical MRI scans and better visualized with ultra-high field MRI (Madsen et al., 2021).

Sensorimotor impairments are among the most common and disabling clinical manifestations (Kister et al., 2013). Transcranial magnetic stimulation (TMS) of the primary motor cortex (M1) provides complementary physiological markers of motor system integrity capturing both corticospinal conduction and interhemispheric (transcallosal) interactions (Vucic and Kiernan, 2017; Madsen et al., 2022). In MS, delayed latencies of the motor evoked potential (MEP) and prolonged corticomotor conduction time (CMCT) (Snow et al., 2019; Madsen et al., 2022) have traditionally been attributed to damage of the corticospinal spinal tract and spinal grey matter (Smith, 1994; Smith and McDonald, 1999). We recently combined TMS of the hand representation in M1 with 7T MRI and showed that the presence of cortical, particularly leukocortical, lesions in the primary sensorimotor hand area (SM1-HAND) contribute significantly to corticomotor conduction slowing (Madsen et al., 2022).

Impaired interhemispheric communication between motor cortices is another hallmark of MS (Wahl et al., 2011; Llufriu et al., 2012), leading to impaired bimanual coordination (Larson et al., 2002) and increased mirror movements (Holzapfel et al., 2024). Yet, the contribution of cortical sensorimotor lesions to direction□specific impairments in interhemispheric motor signaling remains unclear. The ipsilateral silent period (iSP) is a well-established, direction-specific measure of transcallosal inter-hemispheric M1-to-M1 inhibition (Wassermann et al., 1991; Meyer et al., 1995). It reflects the suppression of electromyographic activity in a voluntarily contracting hand muscle evoked by a suprathreshold TMS pulse over the ipsilateral SM1-HAND. Both the latency and magnitude of the iSP are frequently abnormal in MS, and these abnormalities correlate with motor performance and disability (Boroojerdi et al., 1998; Jung et al., 2006; Chieffo et al., 2019). Although diffusion□based measures of callosal microstructure have been linked to iSP alterations (Hoppner et al., 1999; Wahl et al., 2011; Llufriu et al., 2012), it remains unknown whether cortical lesions within M1, either in the stimulated (sending) or inhibited (receiving) hemisphere, contribute to direction□specific disruption of transcallosal inhibition.

In this study we combined ultra-high filed 7T MRI for sensitive detection of cortical lesions, and diffusion-based tractography of transcallosal fibers with single-pulse TMS for measuring bi-directional transcallosal inhibition. We specifically tested whether cortical lesions involving the sensorimotor hand representation impair the transcallosal motor-to-motor inhibitory drive when accounting for callosal microstructural damage.

## 2. Materials and Methods

### 2.1. Participants

We recruited adult people (18-80 years) with relapsing remitting (RRMS) or secondary progressive (SPMS) MS from the outpatient clinic at the Danish MS Center (Copenhagen University Hospital - Rigshospitalet, Copenhagen, Denmark). Exclusion criteria were clinical relapses, corticosteroid therapy or changes in MS-related medication within 3 months of participation, expanded disability status scale (EDSS) above 7, other neurologic or psychiatric disorders, and contraindications to 7T MRI or TMS. Age- and sex-matched healthy controls (HC) were recruited through advertisements. The study cohort has been described previously (Madsen et al., 2022; Madsen et al., 2024). All participants provided written informed consent in accordance with the Declaration of Helsinki. The study was approved by the Committees on Health Research Ethics of the Capital Region of Denmark (H-17033372) and monitored by the local Good Clinical Practice unit, and was preregistered at Clinicaltrials.gov (NCT03653585).

### 2.2. Experimental design

Data presented in the current paper were acquired as part of a larger research project comprising three experimental sessions conducted within a four-week period. During the first session, structural 7T MRI data of the brain were collected, followed by diffusion tensor imaging (DTI) in the second session. Corticomotor physiology was investigated during a third experimental visit. No participants experienced any relapses during their participation.

### 2.3. Clinical examinations

Clinical measures, including EDSS and disease duration, were extracted from clinical records when assessed within three months prior to participation. If no recent evaluation was available, a standardized neurological examination was conducted on experimental day 2. Performance on the 9-hole peg test (9-HPT) was obtained for both hands in all participants.

### 2.4. MRI data acquisition

All MRI data were acquired on a 7T Philips Achieva scanner (Philips healthcare, Best, The Netherlands) equipped with a 32-channel receive head coil (Nova Medical, Wilmington, MA, USA). Structural MRI included T2-weighted turbo spin echo, Magnetization Prepared Rapid Gradient Echo (MPRAGE), Magnetization□Prepared 2 Rapid Acquisition Gradient Echoes (MP2RAGE), and fluid attenuated inversion recovery (FLAIR) sequences. All structural scans were acquired using prospective fat-navigated motion correction (Andersen et al., 2019). A fast T1-weigthed brain scan was obtained for integration with the neuronavigation software. Diffusion weighted imaging (DWI) was based on a multi-slice sequence with 32 diffusion gradient directions. Detailed scan parameters can be found in Supplementary Table 1.

### 2.5. Neurophysiological recordings

#### 2.5.1. Neuronavigated TMS data acquisition

TMS was delivered through an MC-B70 figure-of-eight coil (70mm diameter) connected to a MagPro x100 option stimulator (MagVenture, Skovlunde, Denmark). Surface electromyography (EMG) was recorded bilaterally from the first dorsal interosseous (FDI) muscle using EMG electrodes (Neuroline 700, AMBU, Ballerup, Denmark) and a belly-to-tendon montage. The analog EMG signal was band□pass filtered (5–2000□Hz), amplified (×200–1000) using a D360 amplifier, and digitized at 5000□Hz using Signal 4 software and a 1401 micro Mk□II interface for offline analysis (Cambridge Electronic Design, Cambridge, UK). TMS was applied over the SM1-HAND with the coil placed tangentially to the scalp and the handle oriented approximately 45° to the mid-sagittal plane. Biphasic pulses inducing an anterior-to-posterior followed by a posterior-to-anterior current in the brain were delivered at 0.25Hz with 25% temporal jitter. The individual FDI hotspot was identified using a systematic mini-mapping procedure, selecting the coil position that elicited the largest and most consistent MEPs. Hotspots were marked on individual T1-weighted MRI images using stereotactic neuronavigation (Localite, Skt. Augustin, Germany) to ensure correct and stable coil placement throughout the experiment. Resting motor thresholds (RMT) were determined to the nearest 1% of the maximum stimulator output using the Rossini method (Rossini et al., 2015). EMG activity was constantly monitored, and participants were prompted to relax whenever unintended background contraction was observed.

#### 2.5.2. Corticomotor latency

The corticomotor latency (CML) was assessed while participants performed an isometric contraction of 10% maximal voluntary contraction (MVC). MVC was quantified using a custom-built force transducer and defined as the highest plateau of force production from index finger abduction measured over two trials. During tonic 10% MVC, we applied 11 TMS pulses over the contralateral FDI hotspot at 140% RMT. Participants received continuous visual feedback of their force output and were instructed to maintain the target contraction and disregard TMS□induced perturbations.

#### 2.5.3. Ipsilateral silent period

The iSP was measured to assess directional transcallosal conduction and inhibition between both SM1-HAND. Participants performed an isometric contraction at 80% MVC with their index finger while receiving a 140% RMT TMS pulse over the FDI hotspot ipsilateral to the contracting hand. Fifteen trials were recorded at an interstimulus interval (ISI) of 6 seconds. Participants were instructed to completely relax between trials.

### 2.6. Data analysis

#### 2.6.1. MRI data analysis

All structural images were bias-field corrected, resampled to an isotropic voxel size of 0.5 mm^3^, aligned to MNI space, and co-registered to the MPRAGE image. Volumetric segmentation and surface reconstruction were performed using *FreeSurfer* software (version 7.1.1). Lesion filling was applied, and manual corrections were made when necessary. Cortical grey-matter and subcortical white matter lesions were manually marked by three independent readers using FLAIR, T2-weighted and MPRAGE images. Cortical lesions were classified into type-I (leukocortical), type-II (intracortical), and type III/IV (subpial) subtypes (Bo et al., 2003).

To investigate the local effect of cortical lesions on transcallosal conduction, the bi-lateral SM1-HAND regions served as our regions of interest (ROI). The sensorimotor hand knob (Yousry et al., 1997) was identified by an experienced reader, and a 30-mm spherical ROI was centered on the precentral hand knob, covering both, pre-and postcentral cortices. To examine the impact of cortical lesions on TMS outcome measures, patients were categorized into CL+ (> 1 cortical lesion within the SM1-HAND ROI) and CL- (no cortical lesions within the SM1-HAND ROI). A detailed description of the MRI processing pipeline is provided in Madsen et al. (2022).

#### 2.6.2. Diffusion weighted imaging analysis

DWI data were processed using FSL (Jenkinson et al., 2012) and MRtrix3 (Tournier et al., 2019). Preprocessing followed the procedure described in Madsen et al. (2022). Briefly, images were denoised and corrected for susceptibility□related distortions, eddy-currents and slice-volume motion. Fiber orientation distributions (FOD)s were estimated using constrained spherical deconvolution. Using the FODs, probabilistic tractography was performed between the bilateral SM1-HAND ROIs in MRtrix3, seeding 1,000,000 streamlines in each direction with a step length of 0.18 mm. To ensure anatomically meaningful streamline estimation, the FreeSurfer corpus callosum segmentation was used as an additional inclusion mask. The mask was dilated by one voxel to minimize influence from minor segmentation errors. Tracking was terminated, if FOD was <0.06, deviation angle was >35° or fiber length was <50mm.

Bidirectional tractograms were thresholded at 5% overlap, merged, and binarized to generate the final SM1-HAND to SM1-HAND trans-callosal tract mask. Clear tractography errors were manually corrected. The final mask was used to extract tract-specific white matter lesion volume, total tract volume, mean diffusivity (MD) and fractional anisotropy (FA). MD was calculated after removal of lesioned voxels to avoid bias introduced by pathology (Werring et al., 1999). FA was estimated only from the callosal segment of the transcallosal tract, to avoid cross-fibre regions where FA cannot be clearly resolved with standard DTI models (Wahl et al., 2007; Andersen et al., 2020).

#### 2.6.3. Neurophysiological data analysis

##### 2.6.3.1. Preprocessing

All TMS trials were visually inspected to verify stable force production during stimulation, and trials showing deviations were discarded. Direct current offsets were removed by subtracting the mean pre-stimulus EMG signal from each trace.

##### 2.6.3.2. Corticomotor latency

CML was automatically extracted from the mean rectified EMG trace of the remaining trials. CML was defined as the first local minimum preceding a deflection that exceeded the mean pre-stimulus EMG signal plus three standard deviations. All automatically detected latencies were visually reviewed, and corrected if clear detection errors were identified.

##### 2.6.3.3. Ipsilateral silent period

The iSP onset and offset were automatically detected using a custom Matlab script operating on the mean rectified EMG signal. The iSP onset was defined as the time-point where the EMG signal crossed the mean pre-stimulus EMG level followed by a period of EMG activity below the mean pre-stimulus EMG level minus one standard deviation for at least 5 ms. The iSP offset was determined as the first time point after the suppression period where the EMG signal crossed back above the mean pre-stimulus EMG level and remained above the mean pre-stimulus EMG level plus one SD for at least 5 ms (Chen et al., 2003; Wiemann et al., 2026). All iSPs were visually inspected and manually corrected if the algorithm misidentified boundaries and iSPs with a duration <10ms or an onset <25ms were discarded.

Ipsilateral MEPs (iMEPs) were automatically detected using the same criteria, but applying the threshold of mean pre-stimulus EMG plus one standard deviation. If an iMEP was detected, all individual trials were re-inspected. Trials clearly containing iMEPs (maximum three/15 trials) were removed, and the iSP analysis was repeated. Recordings in which no valid iSP could be identified, or where iMEPs persisted after re□analysis, were excluded (n=8). In addition to iSP onset and duration, normalized iSP depth was quantified as (EMG_pre-stim_-EMG_iSP_)/EMG_pre-stim_ * 100, where EMG_pre-stim_ denotes the mean rectified EMG signal prior to the TMS stimulation and EMG_iSP_ is the mean rectified EMG signal during the iSP period. To account for between-patient variations in ipsilateral corticomotor conduction delay, we also calculated the transcallosal conduction time (TCT) as iSP onset – CML_ipsi_, where CML_ipsi_ denotes the CML of the hand ipsilateral to the stimulated hemisphere.

#### 2.6.4. Data transformations

Variables that were non-normally distributed were log-transformed. For iSP latency, CML and 9-HPT data, substantial right□skewness persisted despite log-transformation. Therefore, these variables were Box-Cox transformed using an optimized lambda values (Osborne, 2010).

### 2.7. Statistical analysis

Demographic statistics were assessed with linear models, Kruskal-Wallis, Chi-square and Mann-Whitney U tests as appropriate. Post-hoc multiple comparison p-values for analyses involving more than two groups were corrected using the *Holm* method.

The relationship between white matter tract pathology (tract volume, lesion volume, MD, and FA) and iSP measures was assessed using Pearson’s correlations between the mean iSP measure (averaged across hands) and each tract specific metric. Correlations were performed in patients only, and P-values were corrected for multiple comparisons within each variable using the *Holm* method (Hothorn et al., 2008).

The origin of the iSP was assessed using a linear mixed-effects model with iSP latency as the dependent variable. Fixed effects included group (HC, RRMS and SPMS) and CML of both the ipsilateral and contralateral hand. Participant was included as a random effect with random intercepts to account for right/left hand observations within individuals.

Additional iSP measures (iSP latency, TCT, iSP duration, and iSP depth) were compared between groups using mixed-effects linear models with participant as random factor with random intercepts and age, sex and hand dominance as covariates of no interest. Exploratory analyses of cortical lesion-type specific effects used the same model structure, but with lesion-type groups (presence/absence of type I, II, III/IV lesions) as fixed factors.

To examine whether additional MRI-based pathology contributed to iSP measures, backwards stepwise linear mixed-effects models were conducted in patients only. Models included group (CL+ and CL-), transcallosal tract volume, MD, FA, and white matter lesion volume in the transcallosal tract as candidate predictors.

Associations between cortical lesions and transcallosal tract pathology was assessed using linear regression models with lesion-presence groups in *either* SM1-HAND ROI as the predictor and age and sex as covariates of no interest. Sensitivity analyses additionally controlled for white matter lesion volume within the tract.

To assess the behavioral impact of iSP metrics, 9-HPT performance was modelled using mixed linear models, including group (HC, CL-, CL+), TCT, iSP duration, and iSP depth from both the ipsilateral and contralateral hand. Finally, relationships between mean iSP measures (averaged across hands) and EDSS were examined using Pearson’s correlations with Holm correction for multiple comparisons.

## 3. Results

### 3.1. Participant demographics

Data were collected from 20 HCs (43.2 ± 13 years, 14 females [70%]), 27 RRMS patients (42 ± 11 years, 19 females [70.4%], median EDSS = 3) and 11 SPMS patients (56 ± 7 years, 7 females [63.6%], median EDSS = 6). Demographic characteristics and MRI summary statistics for healthy controls and patients are shown in Table 1.

**Table 1.**
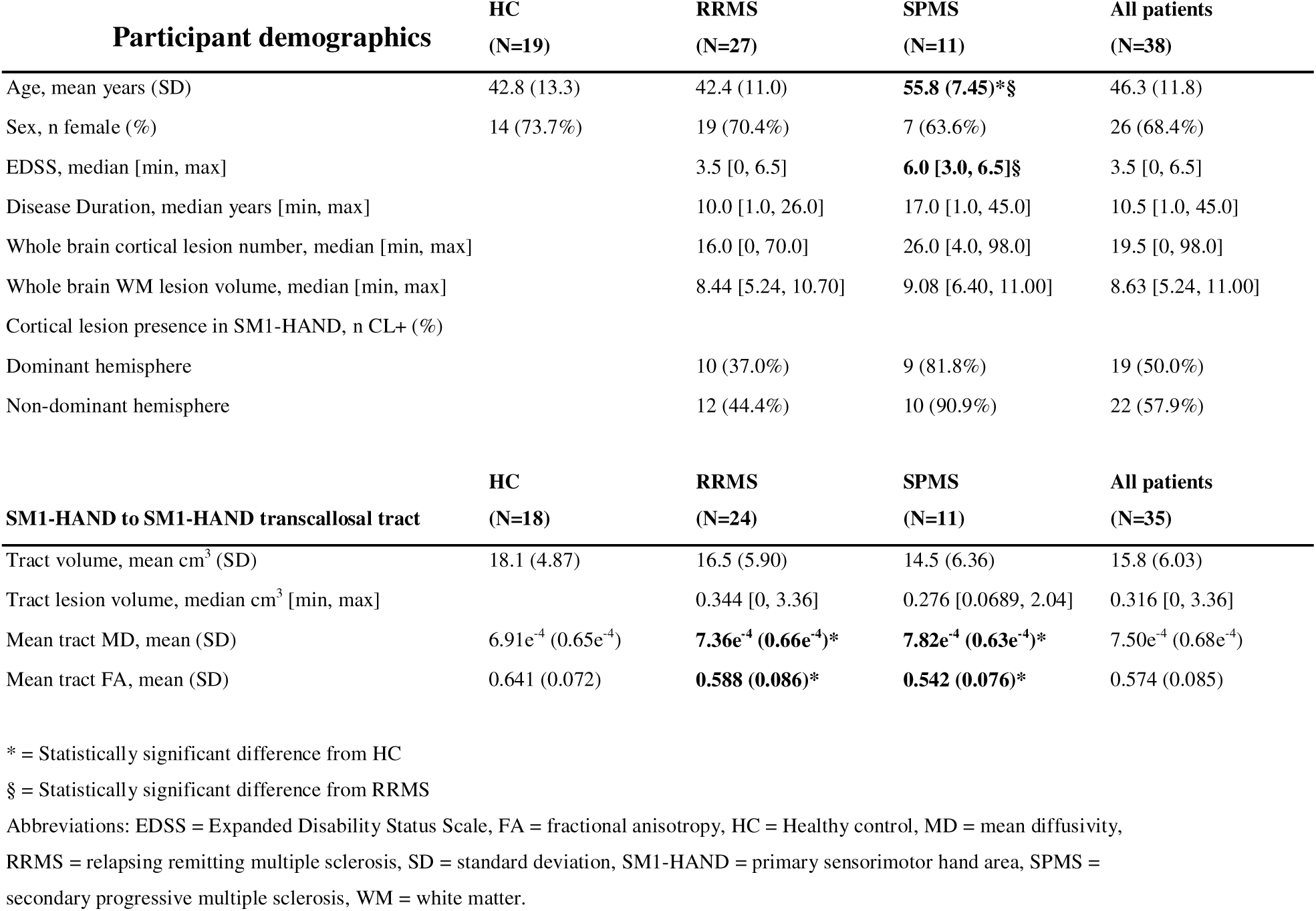
Participant demographics.

Two healthy participants showed iMEPs (one bi-laterally). In five participants (1 HC, 2 RRMS, 2 SPMS; 4.4%) valid iSPs could not be identified in the dominant hand. Thus, eight observations were discarded, leaving 36 HC, 52 RRMS and 20 SPMS observations for directional interhemispheric analysis of transcallosal inhibition. Further, DWI data from one HC and three RRMS patients were excluded due to insufficient quality leaving 18 HCs, 24 RRMS and 11 SPMS patients for tract-specific analyses.

Among the 72 remaining patient hemispheres, 38 (53%) contained one or more cortical lesions within the SM1-HAND ROI and were classified as CL+. Group comparisons showed that SPMS patients were older and had higher EDSS than RRMS patients (P<0.05). Moreover, MS patients exhibited higher MD and lower FA within the SM1-HAND to SM1-HAND transcallosal tract compared to the HC group.

### 3.2. iSP latency is dependent on the central motor latency of the ipsilateral hand

Although the precise neurophysiological origin of the iSP remains to be clarified, converging evidence indicates that the iSP is mediated by homologous transcallosal connections from the stimulated to the unstimulated hemisphere (Meyer et al., 1995; Boroojerdi et al., 1996). The large hemispheric differences in transcallosal conduction latencies often observed in MS provide a suitable context to further substantiate this hypothesis. In our data, CML of the hand muscle ipsilateral to the stimulated hemisphere showed a robust positive linear relation with iSP latency (estimate 0.26 (CI: 0.19-0.33), P<0.001, Fig. 1A), while there was no association between the CML of the contralateral hand muscle and iSP latency (estimate 0.04 (CI: -0.03 – 0.11), P=0.31, Fig. 1B). This pattern demonstrates that the MS-related variation in iSP timing is heavily influenced by the conduction delay of the ipsilateral corticospinal system rather than global motor conduction speed, strongly supporting a transcallosal origin of the iSP.

**Figure 1.**
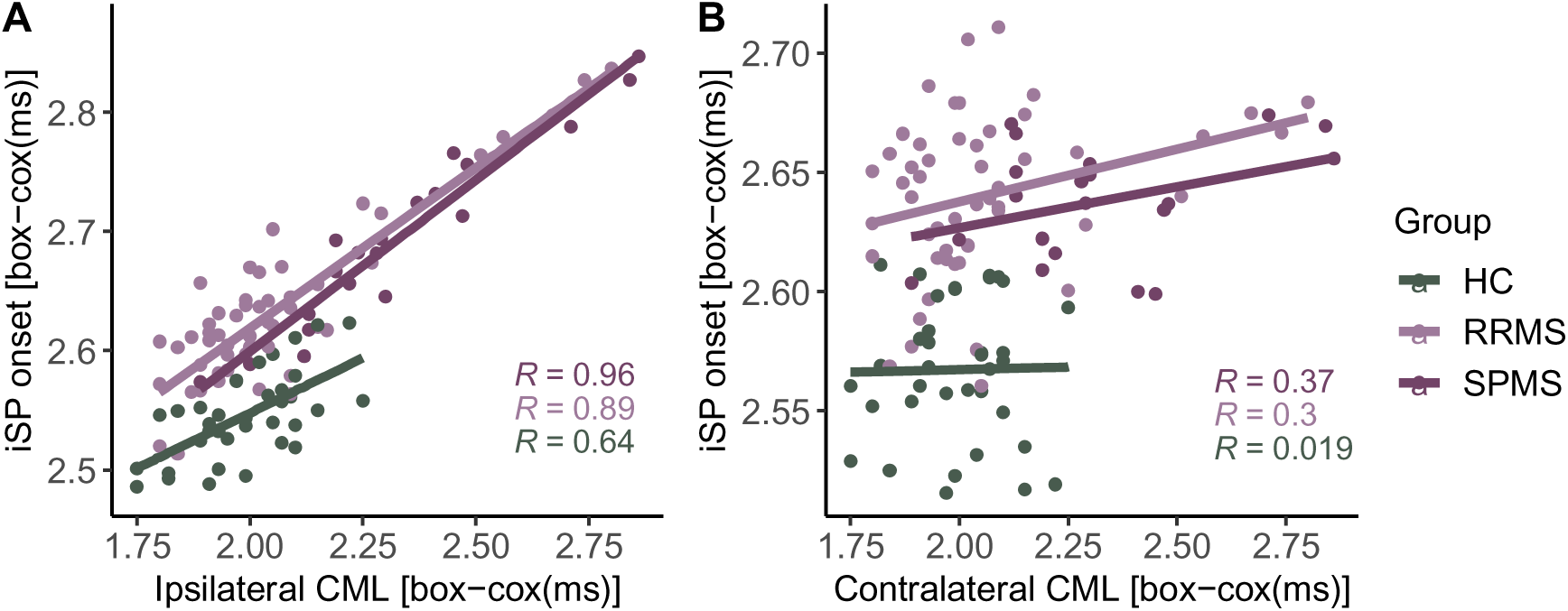
Relationships between CML and iSP latency. **A)** iSP onset (y-axis) plotted against partial residuals of the CML of the hand ipsilateral to the iSP stimulation (x-axis). **B)** iSP onset plotted against partial residuals of the CML of the hand contralateral to the iSP stimulation. Abbreviations: iSP = ipsilateral silent period, CML = corticomotor latency, HC = healthy control, RRMS = relapsing remitting multiple sclerosis, SPMS = secondary progressive multiple sclerosis.

### 3.3. Relationship between presence of cortical lesions and transcallosal conduction

The linear mixed model revealed a group effect on iSP latency (P<0.001). Pairwise comparisons revealed a stepwise pattern. Patients without cortical lesions in the inhibited SM1-HAND (CL- group) region already exhibited longer iSP latencies relative to the HC group (estimate ±standard error. 0.075 ±0.024 box-cox(ms), P=0.004). This MS-related delay was even more pronounced in patients with cortical lesions in the inhibited hemisphere (CL+ group), who showed significantly longer iSP latencies relative to the HC group (0.135 ±0.024 box-cox(ms), P<0.001) and CL- patients (0.06 ±0.018 box-cox(ms), P=0.002, Fig. 2A). A similar pattern emerged when examining the speed of transcallosal conduction. Using the TCT as explanatory variable, we again found a main effect of *group* (P<0.001). Post hoc testing showed a progressive increase in TCT from HCs to CL- patients (2.45 ±0.88 ms, P=0.014) to CL+ patients (vs HC: 4.45 ±0.89 ms, P<0.001, vs. CL-: 1.99 ±0.77 ms, P=0.026, Fig. 2B). Together, these findings indicate that conduction slowing worsen as cortical pathology extends into the inhibited SM1□HAND region.

**Figure 2.**
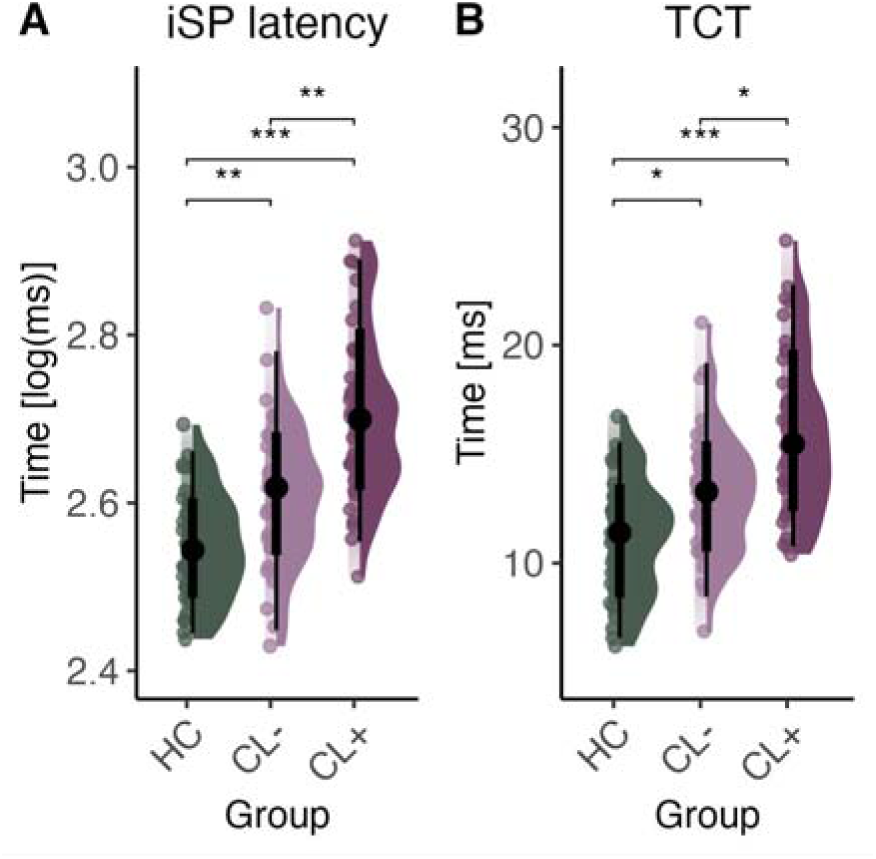
Cortical lesions in the SM1-HAND are associated with transcallosal conduction delays. The figure shows marginal effects and partial residual plots from the linear mixed-effects models of **A)** iSP latency and **B)** TCT. * = P<0.05, ** = P<0.01, *** = P<0.001. CL− = no presence of a cortical lesion in the SM1-HAND, CL+ = presence of a cortical lesion in the SM1-HAND, HC = healthy control, iSP = ipsilateral silent period, SM1-HAND = primary sensorimotor hand area, TCT = transcallosal conduction time.

The presence of cortical lesions could theoretically influence the iSP through either the stimulated or the inhibited cortex. Therefore, we tested for lesion presence in only the stimulated hemisphere as the grouping variable in a similar model. In this case, we found no association between cortical lesion presence and conduction times (iSP latency: P=0.95, TCT: P=0.5). This hemispheric dissociation suggests that conduction delays arise specifically from lesions in the inhibited hemisphere that generates the iSP. Exploratory analyses further identified intracortical type II lesions within the inhibited SM1-HAND region as the primary contributors to this effect (estimate: 2.88 ±0.74 ms, P<0.001). To test whether these effects might be driven by white matter pathology, we conducted a patient-only stepwise linear mixed-effects sensitivity analysis. The only explanatory factor retained was the presence of cortical lesions in the inhibited SM1-HAND (P=0.015). In line with this finding, neither focal nor diffuse white matter pathology showed isolated correlations with any mean iSP measures (P>0.05, Fig. 3). Moreover, we found no associations between cortical or white matter pathology within the transcallosal motor system and iSP magnitude measures such as depth or duration (P>0.05, Fig. 4). Taken together, these results converge on a consistent interpretation, indicating that cortical lesions, particularly intracortical type□II lesions, within the inhibited SM1□HAND region drive transcallosal conduction slowing in the MS group, without a consistent effect of accompanying white□matter pathology.

**Figure 3.**
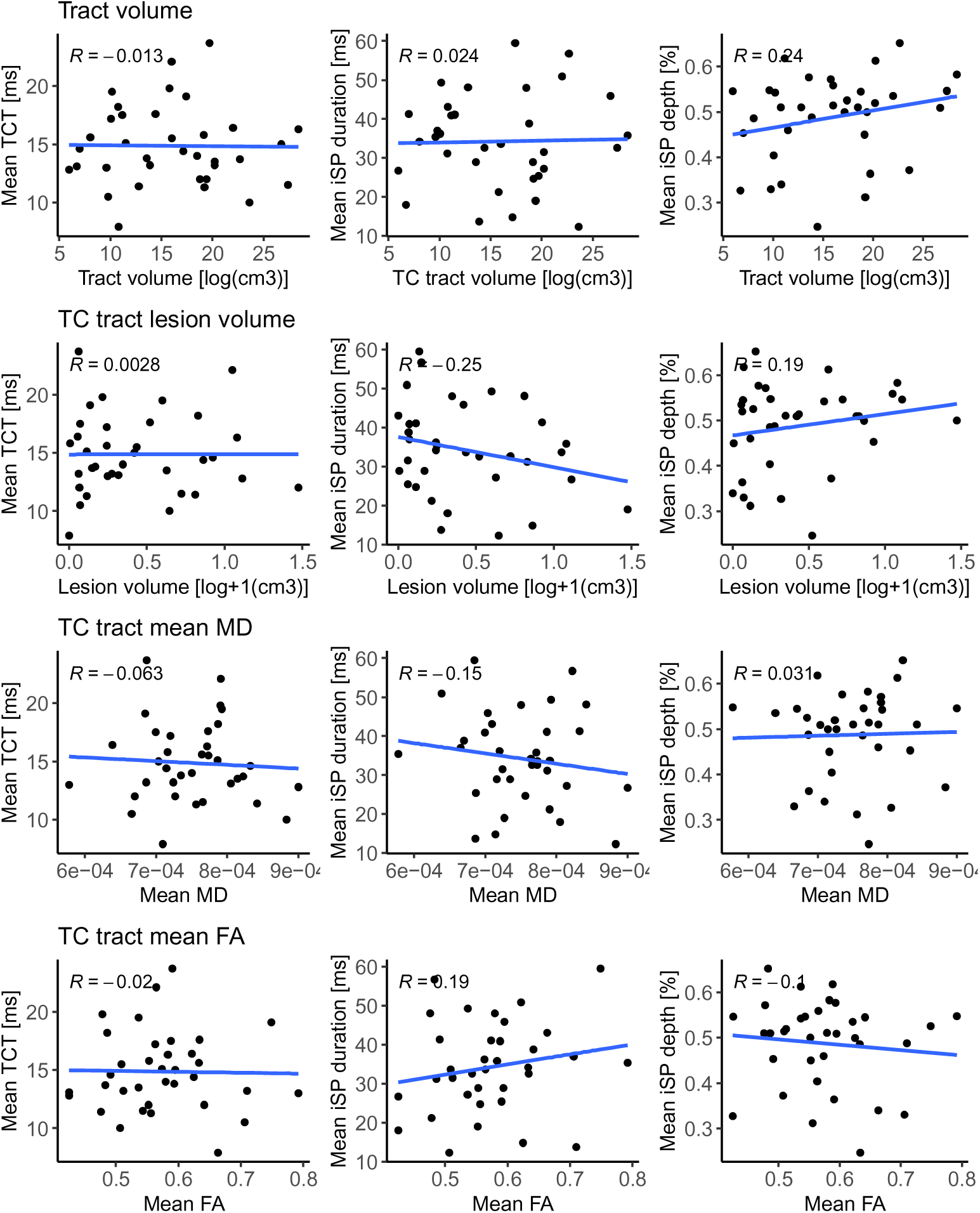
Pearson’s correlations between mean iSP metrics and tract specific white matter pathology. Abbreviations: TCT = transcallosal conduction time, iSP = ipsilateral silent period, TC = transcallosal, MD = mean diffusivity, FA = fractional anisotropy

**Figure 4.**
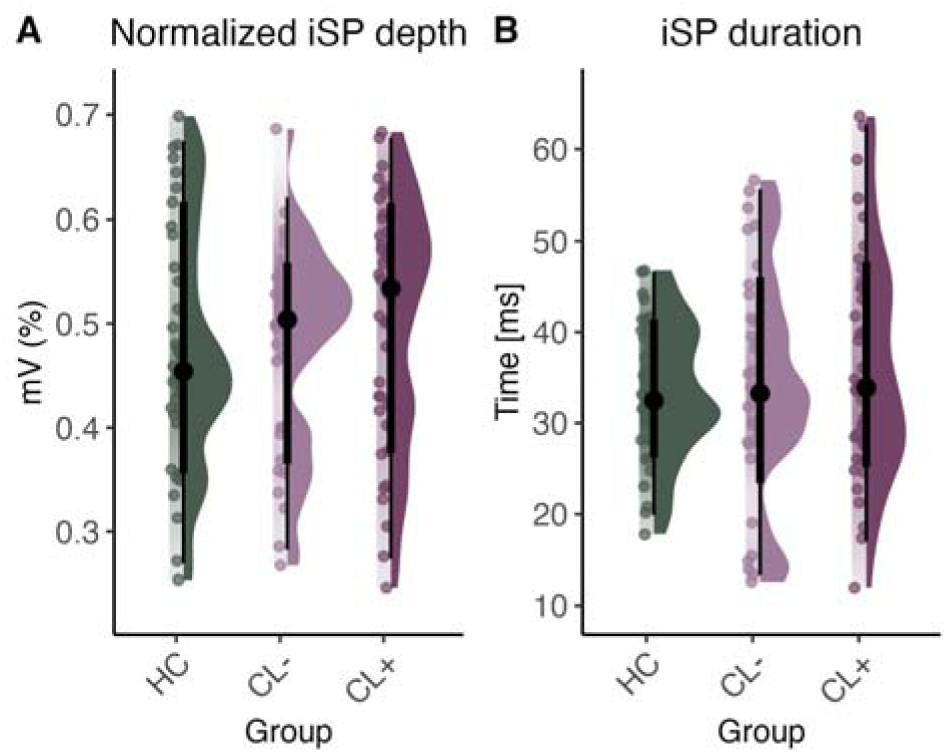
Cortical lesions in the SM1-HAND are not associated with iSP magnitude. The figure shows marginal effects and partial residual plots from the linear mixed-effects models of **A)** normalized iSP depth and **B)** iSP duration. Abbreviations: iSP = ipsilateral silent period, HC = healthy control, CL- = no cortical lesion presence in the SM1-HAND area, CL+ = cortical lesion presence in the SM1-HAND area.

### 3.4. Associations between SM1-HAND cortical lesions and transcallosal tract pathology

It remains an open question whether cortical lesions are accompanied by increased pathology in the white matter tracts they are connected to. To address this, we examined whether structural properties of the transcallosal SM1–HAND tract, such as volume, FA, MD and tract-specific white matter lesion volume, were associated with the presence of SM1-HAND cortical lesions. Patients were grouped according to whether either SM1-HAND region contained a cortical lesion (CL+ =24, CL- =14). Group status was neither associated with differences in normalized tract volume (P=0.11) nor tract-specific white matter lesion volume (P=0.67, only patients included), but with higher mean MD of the transcallosal motor tract volume (P=0.006), being higher for CL+ patients (7.25e^-5^ ±2.15e^-5^ mm^s^/s, P=0.004, Fig. 5B) compared to the HC group. We also found a significant group effect on mean FA in the transcallosal motor tract volume (P=0.025) with CL+ patients having lower FA than HCs (-0.073 ±0.026, P=0.019, Fig. 5C). These effects lost significance after controlling for white matter lesion volume within the transcallosal tract (MD: P=0.072, FA: P=0.48), indicating that MS-related white□matter injury rather than cortical lesions accounts for these differences in transcallosal white mater microstructure.

**Figure 5.**
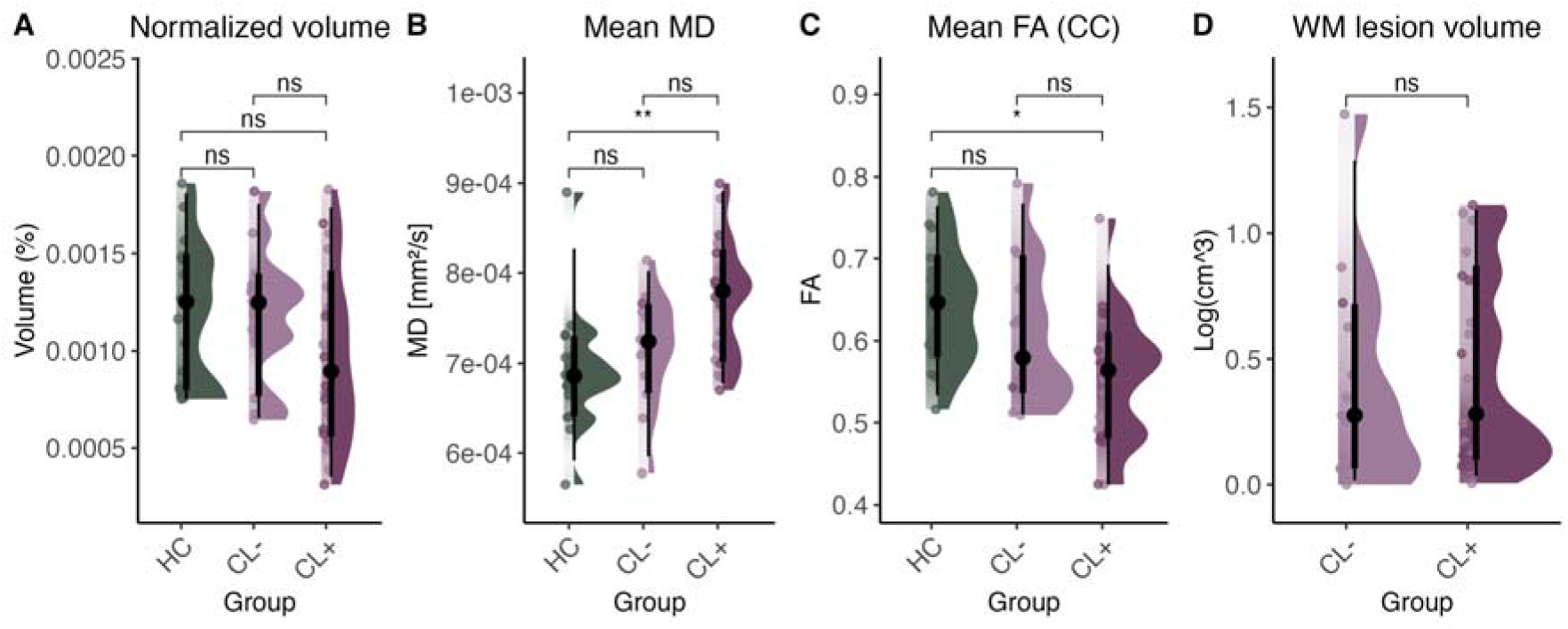
Relation between cortical lesion presence and downstream tract pathology. Point and density plots of **A)** tract volume normalized to intracranial volume, **B)** Mean MD from the SM1-HAND to SM1-HAND transcallosal tract (white matter lesions excluded), **C)** Mean FA from the callosal part of the SM1-HAND to SM1-HAND transcallosal tract and **D)** White matter lesion volume of the SM1-HAND to SM1-HAND transcallosal tract. Abbreviations: SM1-HAND = primary sensorimotor hand area, MD = mean diffusivity, FA = Fractional Anisotropy, WM = white matter, ns = not significant, HC = healthy control, CL- = no cortical lesion presence in the SM1-HAND area, CL+ = cortical lesion presence in the SM1-HAND area.

### 3.5. Correlations with clinical impairment

We next examined the clinical correlates of iSP measures. None of the examined iSP measures correlated with EDSS (P>0.05). However, exploratory analyses revealed that performance on the 9-HPT was linked to iSP duration of the contralateral hand (P=0.007) independent of group status (HC, CL- and CL+, Fig. 6B). This suggests that transcallosal inhibitory function may have general behavioral relevance for skilled hand performance, independently of the presence of MS-related structural pathology.

**Figure 6.**
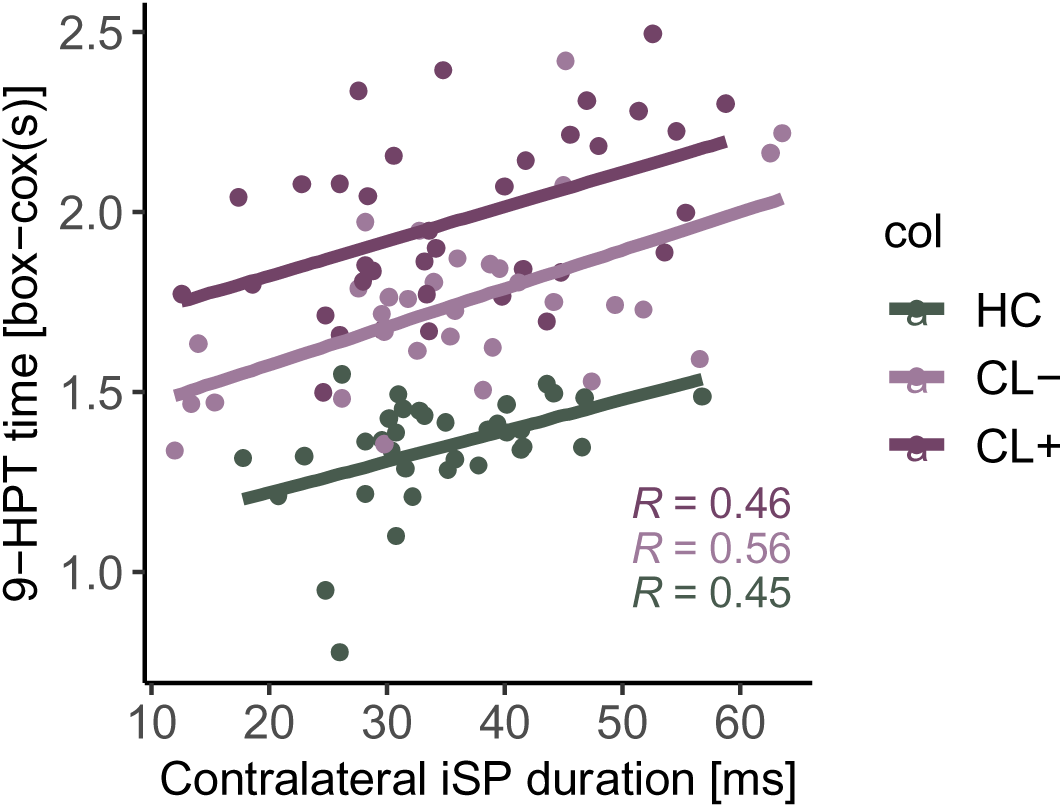
Associations with hand motor function. Scatter plots show the relationship between Box-cox trans-formed time to complete the 9-hole peg test and iSP duration of the contralateral hand. Abbreviations: iSP = ipsilateral silent period, 9-HPT = 9-hole peg test, HC = healthy control, CL- = no cortical lesion presence in the primary sensorimotor hand area, CL+ = cortical lesion presence in the primary sensorimotor hand area.

## 4. Discussion

Using TMS-evoked transcallosal suppression of voluntary activity, we combined focal TMS with 7T MRI to examine how the presence of cortical lesions in the SM1-HAND influences transcallosal inhibitory interactions in people with MS. Patients who had a cortical lesion showed a delayed onset of transcallosal motor inhibition compared with patients without such lesions. This delay was direction-specific: Transcallosal inhibition was selectively slowed when inhibition was directed toward the lesioned hemisphere. Intracortical type□II lesions appeared to drive this effect. By contrast, the delay in TMS-evoked transcallosal interhemispheric inhibition was not related to conventional DTI-based markers of callosal tract integrity, including MD, FA and white matter tract lesion load.

### 4.1. Direction-specific link between cortical lesions and transcallosal conduction

We found a direction-specific delay in the onset of transcallosal motor inhibition. The presence of cortical lesions in SM1-HAND was associated with longer TCT from the stimulated to the lesion bearing (non-stimulated) motor cortex but not vice versa, revealing a delayed build-up of inhibitory influence on the corticospinal output in the lesioned cortex.

We have previously shown with single-pulse TMS that cortical lesions in the SM1-HAND area are associated with delayed cortico-motor conduction time from the lesion bearing motor cortex to the contralateral hand (Madsen et al., 2022). Expanding this finding, the single-pulse based assessment of iSP showed that cortical lesions delay both the processing of incoming transcallosal signals and the outgoing descending corticospinal output. Our exploratory analyses linked type II intracortical lesions to the delay in TCT, while we previously found a coupling between the presence of type I leukocortical lesions and prolonged CMCT (Madsen et al., 2022). Together, these findings suggest that intracortical lesions may preferentially affect cortical circuits that process axonal inputs from other cortical areas, while leukocortical lesions rather affect cortical circuits generating descending pyramidal tract motor output.

Because our previous spectroscopic and structural analyses had shown that cortical lesions affect the local excitation/inhibition balance (Madsen et al., 2024) and corticomotor conduction to the contralateral hand (Madsen et al., 2022), we hypothesized that cortical lesions not only affect the onset but also the magntiude of transcallosal motor inhibition. Indeed, an animal model of cortical and callosal MS pathology was associated with trans-callosal conduction delays and a reduced amplitude of the transcallosal compound action potential (Mangiardi et al., 2011). A more recent study showed the cortical demyelination in Cuprizone fed mice was associated with millisecond delays in conduction latency along with reduced temporal precision of long-range signals (Jamann et al., 2025). However, most patient-related studies have failed to detect any differences in the magnitude of transcallosal motor inhibition between MS patients and HCs (Boroojerdi et al., 1998; Hoppner et al., 1999; Jung et al., 2006; Holzapfel et al., 2024). Moreover, iSP duration in MS has been found to be either longer (Boroojerdi et al., 1998; Hoppner et al., 1999; Jung et al., 2006), unaffected (Llufriu et al., 2012) or even shorter than in HCs (Lenzi et al., 2007). Accordingly, we found no difference in the depth or duration of iSP between MS patients and HCs or between hemispheres with or without a cortical lesion. One reason for these discrepant findings could be that measures of iSP magnitude are highly dependent on methodological choices such as stimulation intensity and are affected non-linearly by pre-stimulation EMG magnitude (Wiemann et al., 2026). It is possible that methodological factors also influenced the sensitivity of our measures to detect potential differences between groups, especially given that we used a rather high pre-stimulus contraction level (80% MVC).

### 4.2. Transcallosal white matter pathology and interhemispheric conduction

The iSP is thought to reflect cortico-cortical transcallosal inhibition of homologous motor representations in M1. This notion was driven by findings that patients with callosal agenesis show no or highly reduced iSP (Meyer et al., 1995), but patients with subcortical stroke and no contralateral MEP sign showed preserved iSP (Boroojerdi et al., 1996). Our analyses support a transcallosal mechanism, as iSP onset latency was only associated with corticomotor conduction speed of the hand ipsilateral but not contralateral to the stimulation. This indicates that the iSP is mediated through the same corticospinal pathway as MEPs evoked in the ipsilateral hand and implies that the corticofugal volley travels along transcallosal interhemispheric connections from the stimulated SM1-HAND to the non-stimulated SM1-HAND.

Although the iSP is likely caused by a transcallosal mechanism, we found no associations between tract-specific measures of white matter lesion load or microstructural integrity and iSP-based metrics reflecting the timing or magnitude of transcallosal motor inhibition. This negative finding aligns with previous studies that failed to show any significant relationships between TCT and MRI measures of MS pathology (Boroojerdi et al., 1998; Jung et al., 2006; Lenzi et al., 2007; Holzapfel et al., 2024). Using a dual-site conditioning-test TMS paradigm, Wahl et al. (2011) assessed the relation between the magnitude of short-latency interhemispheric inhibition and mean FA in the motor portion of the corpus callosum in patients with early RRMS (Wahl et al., 2011). Callosal FA was found to be reduced in patients compared to HCs in the absence of macroscopic MRI lesions.

Importantly, only HCs showed a significant positive linear correlation between callosal FA and the magnitude of short-latency interhemispheric inhibition, while this structure-function relation had vanished in the patient group. Together, these findings indicate that DTI-derived microstructural metrics are sensitive to damage, but lack mechanistic specificity, rendering pathophysiological interpretation ambiguous (Bauer et al., 2020). More advanced diffusion-based acquisition and analyses techniques might be more powerful in linking transcallosal pathophysiology to transcallosal microstructural change (Andersen et al., 2020; Skoven et al., 2025). Regardless of the diffusion weighted MRI data acquisition and modelling approach chosen, diffusion-based MRI read-outs cannot resolve microstructural changes caused by damage of left-to-right or right-to-left transcallosal projection. Here, TMS of the motor cortex offers a powerful complementary means to reveal how MS or other brain diseases affect interhemispheric pathways and function, reflecting the speed, magnitude, and direction of transcallosal conduction.

### 4.3. Cortical lesions and white matter tract damage

Previous studies have shown that cortical lesions are associated with cortical cell body loss (Vercellino et al., 2005; Granberg et al., 2017; Krijnen et al., 2025), and a reduction in N-acetylaspartate a marker of neuronal integrity (Madsen et al., 2024). Moreover, cortical thinning of the primary motor cortex has been related to corticospinal tract integrity (Bergsland et al., 2015). However, while diffusion metrics have been shown to be altered in cortical lesions (Poonawalla et al., 2008; Calabrese et al., 2011; Granberg et al., 2017) the relationship to pathology of white matter tracts does not seem to be spatially specific (Louapre et al., 2016).

Correspondingly, we found that the impact of cortical lesions on transcallosal MD or FA was dissociated and driven by co-existing white matter damage. In addition, the SM1-HAND ROI used in our study most likely also contain fibers in corticospinal and cortico-thalamic tracts thereby decreasing specificity to transcallosal pathology. The exact relationship between cortical and transcallosal microstructural damage needs to be disentangled in future studies.

### 4.4. Limitations

Our study has several limitations. The cross-sectional study design limits the causal interpretations of our results, making it difficult to assess the temporal associations between lesion formation, degeneration, potential remyelination and our iSP measures. Moreover, even though 7T MRI shows substantial improvement in cortical lesion detection compared to 3T, it still catches only a fraction of cortical pathology in MS patients (Madsen et al., 2021). Moreover, MS is also associated with changes in normal appearing grey matter (Cercignani et al., 2001), which we did not take into consideration in the current study. Lastly, transcallosal function was assessed with single-site, single-pulse TMS during tonic contraction. We did not assess transcallosal motor inhibition with dual-site conditioning-test approach which would have enabled comparisons at rest and during contraction (Ferbert et al., 1992; Wahl et al., 2007). We also did not assess dynamic changes in transcallosal motor inhibition during bimanual motor tasks, neither did we assess bimanual motor competencies in our patients. Our findings motivate to study the impact of cortical lesions on these aspects of transcallosal motor control in future studies.

## Conclusions

Our findings provide new insight into the cortical contribution to disrupted interhemispheric motor communication in people with MS. We show that the presence of cortical lesions in the inhibited SM1-HAND area is associated with a directional delay in conduction latency of the transcallosal system, an effect driven by intracortical type II lesions. The results suggest that intracortical lesions in the receiving SM1-HAND affect the ability of regional circuits to rapidly translate incoming transcallosal volleys into inhibitory activity that can successfully shut down tonic voluntary output of the contralateral hand.

## Data Availability

Pseudonymized data can only be shared with a formal Data Processing Agreement and a formal approval by the Danish Data Protection Agency in line with the requirements of the GDPR.

## Author Contributions

MAJM: Conceptualization, Formal analysis, Funding acquisition, Data collection, Formal Analysis, Visualization, Writing – original draft. LC: Methodology, Formal Analysis, Writing – original draft. VW: Methodology, Formal Analysis, Writing – review and editing. HL: Methodology, Formal Analysis, Writing – review and editing. JRC: Data curation, Writing – review and editing, MB: Data Curation, Writing – review and editing. FS: Data Curation, Funding Acquisition, Supervision, Writing – review and editing. HRS: Conceptualization, Funding Acquisition, Project administration, Supervision, Writing – review and editing.

## Glossary

9-HPT: 9-hole peg test
CL+: Cortical lesion presence in SM1-HAND
CL-: No cortical lesion presence in SM1-HAND
CML: Corticomotor latency
CMCT: Corticomotor conduction time
DTI: Diffusion tensor imaging
DWI: Diffusion weighted imaging
EDSS: Expanded disability status scale
EMG: Electromyography
FA: fractional anisotropy
FDI: First dorsal interosseous
FLAIR: fluid attenuated inversion recovery
FOD: Fiber orientation distribution
HC: Healthy control
iMEP: Ipsilateral motor evoked potential
iSP: Ipsilateral silent period
ISI: Interstimulus interval
M1: Primary motor cortex
MD: Mean diffusivity
MEP: Motor evoked potential
MP2RAGE: Magnetization prepared 2 rapid gradient echo
MPRAGE: Magnetization prepared rapid gradient echo
MRI: Magnetic resonance imaging
MS: Multiple sclerosis
MVC: Maximal voluntary contraction
RMT: Resting motor threshold
ROI: Region of interest
RRMS: Relapsing remitting multiple sclerosis
SM1-HAND: primary sensorimotor hand area
SPMS: Secondary progressive multiple sclerosis
TCT: Transcallosal conduction time
TMS: Transcranial magnetic stimulation

