## Supplemental Table 1 for "Delayed Transcallosal Conduction to the Lesioned Sensorimotor Cortex in Multiple Sclerosis: A combined TMS 7T-MRI Study"

**Supplementary Material**

**Supplementary table 1. Summary of MRI acquisition parameters.**

^a^All 3D data were collected with sagittal read-out,

^b^Multi-slice DTI data were acquired axially.

^c^ The MPRAGE echo spacing/RAGE-train TR was 7.5ms.

^d^The MP2RAGE echo spacing/RAGE-train TR was 8ms, and data were acquired with 0.775 halfscan along y, TFE factor 128.

^e^The T2w-TSE factor was 200.

^f^The DTI acquisition used a single b-value of 1000 s/mm^2^ over 32 gradient diffusion encoding directions.

Abbreviations: FOV = field-of-view, rec = reconstructed, TE = echo time, TR = repetition time, TI = inversion time, FA = flip angle, MoCo = prospective motion correction, SENSE = sensitivity encoding (parallel acceleration factors), AP = anterior-posterior, RL = right-left, iso = isotropic, MS = multi-slice, EPI = echo planar imaging,

| **Sequence** | **FOV**  **[mm]** | **Voxel size [mm]** | **Rec voxel size [mm]** | **TE**  **[ms]** | **TR**  **[ms]** | **TI**  **[s]** | **FA**  **[°]** | **MoCO** | **SENSE** | **Scan time** **[min]** |
| --- | --- | --- | --- | --- | --- | --- | --- | --- | --- | --- |
| 3D^a^ MPRAGE^c^ | 228 x 228 x 166 | 0.65 iso | 0.65 iso | 3.2 | 2200 | NA | 8 | Yes | AP: 1.6, RL: 1 | 7:13 |
| 3D MP2RAGE^d^ | 205 x 205 x 192 | 0.85 x 0.85 x 0.8 | 0.8 iso | 3.5 | 5500 | 404/  3004 | 6/5 | Yes | AP: 1.8, RL: 1.8 | 11:39 |
| 3D FLAIR | 230 x 230 x 168 | 0.7 iso | 0.69 x 0.69 x 0.7 | 391 | 5000 | 1.83 | 55 | Yes | AP: 2, RL: 1.5 | 11:10 |
| 3D T2w-TSE^e^ | 256 x 256 x 190 | 0.8 iso | 0.4 iso | 319 | 3719 | NA | 100 | Yes | AP: 2, RL: 2 | 11:54 |
| 3D T1w | 246 x 246 x 174 | 0.99 x 1 x 1 | 0.85 x 0.85 x 1 | 2.2 | 5.0 | NA | 7 | No | AP: 2, RL: 2.5 | 01:55 |
| MS DTI-EPI^b,f^ | 200 x 200 x 121 | 1.79 x 1.85 x 1.8 | 1.79 x 1.79 x 1.8 | 54 | 13935 | NA | 90 | No | AP:4 | 8:08 |
